# Hip fracture rehabilitation in hospital and community settings: A rapid review of the evidence

**DOI:** 10.1101/2025.01.15.25320606

**Authors:** Llinos Haf Spencer, Kodchawan Doungsong, Mohammed Albustami, Shanaz Dorkenoo, Rhiannon Tudor Edwards, Nefyn Williams

## Abstract

**Background:** Proximal femoral or hip fracture is a common problem among older adults. With an increase in patients with hip fracture comes and increase in the number of patients who require post-surgery rehabilitation. Our aim was to review effectiveness of rehabilitation after hip fracture studies.

**Methods:** For this rapid review (RR) four databases, PubMed, PsycInfo, CINAHL and Cochrane Library, were searched for studies published between 1^st^ January 2017 and 15^th^ May 2023. The population were: people with hip fracture. Interventions: any hospital or community-based hip rehabilitation. Comparisons: improvements in function, and the outcomes were: physical function, quality of life and cost.

**Findings:** Six studies were included in this RR which focussed on hip fracture rehabilitation programmes published since 2017. The studies were conducted in North America (USA), Europe (Germany, Austria, and Switzerland) and Asia (Hong Kong and China). Some randomised controlled trials and some cohort studies found beneficial effects of rehabilitation for older people who had undergone hip replacement treatments. The length of follow-up period ranged from 15 days to one year after surgery. Even though psychological distress, including loss of confidence and frustration, was the most important predictor of depression and anxiety in patients with hip fractures, only one study assessed anxiety and depression status of patients after surgery. It was not possible to combine the results in a meta-analysis as all the studies had different follow-up periods and used different outcome measures.

**Interpretation:** The goal of achieving personalised rehabilitation treatment is possible and this concept of goal directed personalised rehabilitation should be developed and economically evaluated further. A strength of this review is that robust, recent, and relevant papers regarding rehabilitation following hip replacement were found and described. However, the main limitations were that none of the included studies conducted a cost-analysis of the intervention that they described, and no UK based study was included.

## Background

With an ageing population, proximal femoral or hip fractures are a major public health issue (Peeters et al., 2016). Hip fractures are a challenging global health problem with significant socioeconomic consequences for the patients and their families and health care budgets (Marques et al., 2015). The average age of hip fracture patients is 83 years old and 79% are female. Comorbid conditions and cognitive impairments are common in this group of patients (Van Balen et al., 2003).

The financial cost of managing hip fracture patients is high. According to recent estimates, the annual NHS inpatient burden of hip fracture was £639 million to £664 million in England and £50 million to £55 million in Wales from 2016/7 to 2018/9 (Baji et al., 2022).These costs are directly proportional to several variables, including the length of hospital stay, timing of surgery, availability of specialised orthopaedic–geriatric units, and access to rehabilitation after the hospitalisation.

Common complications include new falls, pneumonia, and new fractures (Hansson et al., 2015). Following a hip fracture operation, an older person’s recovery is enhanced if they are provided with an optimistic, well-coordinated rehabilitation programme. An early rehabilitation approach should prevent functional decline and complications and preferably include a multidisciplinary team (physiotherapy, occupational therapy, nutrition, social work, psychology, and medicine) with the integration of orthogeriatric and rehabilitation services (Dyer et al., 2021). The rehabilitation team should meet with the patient regularly and provide appropriate treatments based on goals which have been set with the patient and family. Goals should be reviewed and assessed at regular intervals to ensure appropriate ongoing input to maximize recovery (Beer et al., 2022).

Better outcomes regarding mobility is associated with early and high doses of mobility training (Dyer et al., 2021). Rehabilitation programmes should begin soon after hospital admission and continue after hospital discharge (Dyer et al., 2021). Rehabilitation training should include goal-directed mobilisation practice including tailored balance and functional exercises to promote independence (Beer et al., 2022).

In recent years, four systematic reviews have been conducted to investigate outcomes following hip replacements (Alexiou et al., 2018; Fairhall et al., 2022; Handoll et al., 2021; Peeters et al., 2016). A systematic review from 2016 reported on studies measuring health status or health related quality of life (HRQoL) in patients older than 65 years with a hip fracture (Peeters et al., 2016). Forty-nine studies were included which were all either randomised controlled trials (RCTs) or prospective cohort studies. Following hip fracture, physical, social, and emotional functioning were affected. The health status and HRQoL of most patients recovered in the first six months after fracture. However, their health status did not return to pre-fracture levels. Mental state, pre-fracture functioning on physical and psychosocial domains, comorbidity, female gender, nutritional status, postoperative pain, length of hospital stay, and complications were negatively associated with health status or HRQoL (Peeters et al., 2016).

A systematic review from 2018 (Alexiou et al., 2018) included 20 RCTs, and 29 prospective cohort studies. The size of the samples analysed ranged from 61 to 33,152 patients. The age range of the cohorts was 72–84 years, and the length of follow-up ranged from six weeks to four years. For the assessment of the outcomes, the SF-12 and SF-36, Barthel Index, and EQ-5D questionnaire were used (Alexiou et al., 2018). During the first months after a hip fracture, the physical functioning of all the patients was seriously affected, with a subsequent detrimental impact on the health status and HRQoL, both of which showed an incomplete recovery in most patients. Only four of the studies reported a complete recovery based on the pre-fracture levels of HRQoL and health status (Comans et al., 2013; Hansson et al., 2015; Mariconda et al., 2016; Van Balen et al., 2003)

In terms of regaining previous mobility, this ranged from 18% to 29% (Hansson et al., 2015; Van Balen et al., 2003). In addition, most of the greatest recovery takes place within the first six months, while some improvement in the physical status is observed up to a year postoperatively (Alexiou et al., 2018).

The aim of this rapid review was to report on studies which have evaluated the effectiveness or cost-effectiveness of rehabilitation interventions following hip fracture since the previous systematic reviews found from the worldwide literature from 2017.

## Methodology

The protocol for this rapid review was published on PROSPERO (Spencer et al., 2023). Four databases were searched including MEDLINE, PsycINFO, CINAHL and EMBASE. In terms of Population, Interventions, Context and Outcomes (PICO) (Schardt et al., 2007) the primary outcomes included: functional outcomes such as activities of daily living (see Supplementary files 1 and 2 for the PICO table and search strategy via Medline). Measures of effect were reported according to the guidelines of the Cochrane Collaboration to measure the effectiveness of an intervention. Secondary outcomes included: Quality of life outcomes and cost of interventions/programmes.

Systematic Reviews, RCTs, primary studies including cross-sectional, cohort, longitudinal, and case studies/case reports were included. Editorials, letters to editors, conference abstracts, commentaries, and viewpoint papers were excluded.

The PRISMA diagram is shown in Figure 1.

**Figure 1.**
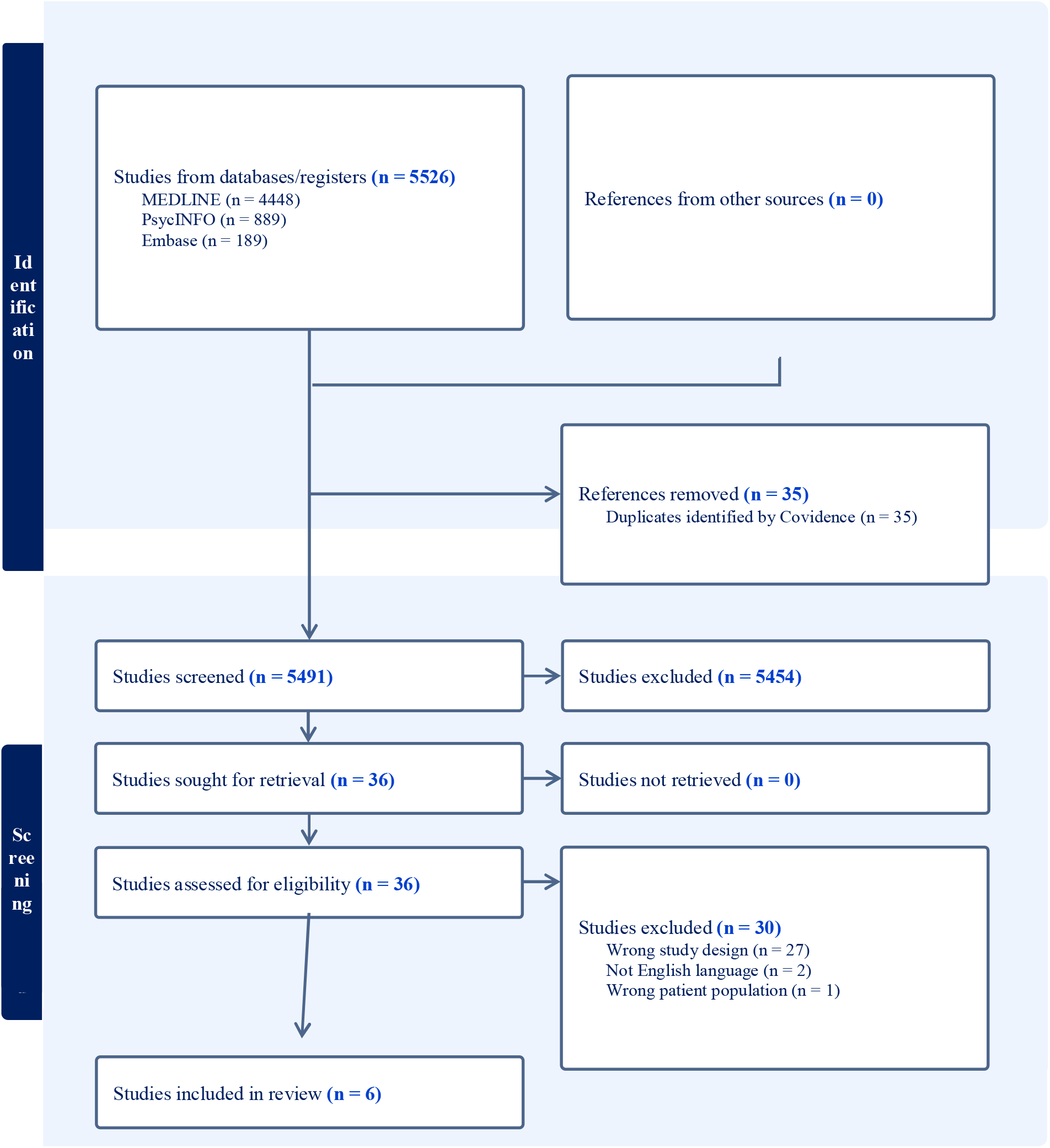
PRISMA 2020 flow diagram of included studies (Page et al., 2021).

### Data extraction methodology

Data extraction was performed by two reviewers (LHS and KD). Data extracted included: study country, study design, intervention type (or usual care), data collection methods, dates, sample size, type of participants (including ethnic groups), primary findings and additional findings.

### Quality appraisal methodology

The quality assessment was undertaken by two reviewers (LHS and KD) with a third reviewer checking a third of the papers for quality assurance purposes (NW). The Joanna Briggs Institute (JBI) critical appraisal tools were used for the quality appraisal of randomised controlled trials, and cohort studies (Moola et al., 2017). (See Supplementary file 3).

## Results

Six studies which matched the inclusion criteria were included in this rapid review. Three studies were randomised controlled trials (RCTs) (Aftab et al., 2020; Magaziner et al., 2019; Zhang et al., 2022) three were cohort studies (Schoeneberg et al., 2021; Xiang et al., 2021; Zhong et al., 2021). See Table 1. Summarised outcome measures that were used in the included studies are shown in Table 2.

**Table 1.** Summary characteristics of included studies.

| **Name and date** | **Country** | **Number of participants in the intervention group** | **Follow-up period** | **Main finding** | **Economic analysis?**<br>**Yes/No** |
| --- | --- | --- | --- | --- | --- |
| Aftab et al., 2020<br>RCT | South Korea | n=20 in the intervention group | Follow-up period: On 2<sup>nd</sup> postoperative day and after 10th FIRM session on 15<sup>th</sup> postoperative day. | The post intervention comparison did not show any significant difference ($p>0.05$) in walking ability, overall activities of daily living and cognitive functioning. | No |
| Magaziner et al., 2019<br>RCT | USA | n=105 in the Training Group arm. | 16 and 40 weeks after randomization. | There was no significant difference in community ambulation between the training group and the active control group ($p = .19$ at 16 weeks and $p=.98$ at 40 weeks). | No |
| Zhang et al., 2022<br>RCT | China | n=29 in the telerehabilitation group. | 1 month and 3 months after surgery | At 1 and 3 months after the intervention, the difference between the intervention and control groups were significant ($p < 0.05$); for patients after hip replacement, the timed up-and-go test (TUG), and short physical performance battery (SPPB) scores in the telerehabilitation group were better than those in the telephone group at 3 months after the intervention, and the difference was significant ($p < 0.05$). | No |
| Schoeneberg et al., 2021<br>Cohort | Germany Switzerland and Austria | n = 7,066 in the Early geriatric rehabilitation (EGR) group. | 120 days post-surgery | The Early geriatric rehabilitation (EGR) showed a positive influence of anti-osteoporotic treatment ($p < 0.001$) and mortality ($p = 0.011$) but led to a slight reduction in QoL ($p = 0.026$). | No |
| Xiang et al., 2021<br>Cohort | China and Hong Kong, China | n = 148 in early mobilization, n= 136 in late mobilization | Patients attended clinic follow-up visits at 6 and 12 weeks. The 1-year visit was conducted via a telephone interview. | At 6 weeks, early mobilization resulted in a significantly better Modified Barthel Index than late mobilization (mean [SD]: 83.7 [12.0] vs. 67.0 [17.5], $p < .001$). Adjusted mixed effects model showed significantly higher Modified Barthel Index for early mobilization at postoperative visit, 6 weeks, and 12 weeks (all $p < .001$). Patients in the early mobilization group had slightly better EQ-5D Index at 12 weeks than patients in the late mobilization group (mean: 0.91 vs 0.87, $p = .002$). | No |
| Zhong et al., 2021<br>Cohort | China | n=180 received rapid rehabilitation nursing | Within one year after surgery. | Compared with the patients in the conventional rehabilitation group, those in the rapid rehabilitation group had shorter hospital stays ($11.5 \pm 1.2$ day vs $15.5 \pm 2.3$ day, $p = .021$), resumed off-bed activities sooner ($20.5 \pm 3.4$ hours vs $61.8 \pm 4.7$ hours, $p = .001$), had less postoperative pain ($4.0 \pm 1.2$ vs $6.5 \pm 1.1$, $p < .001$), and lower anxiety and depression scores (anxiety score: $24.4 \pm 2.1$ vs $47.9 \pm 2.9$; depression score: $25.8 \pm 1.8$ vs $43.7 \pm 1.7$, $p < .001$). | No |

**Table 2.** Summarised outcome measures that used in included studies.

| Categories | Outcome measures |
| --- | --- |
| **Performance test** |  |
| Physical performance | Modified Physical Performance Test (mPPT) |
| Balance | NHATS Balance score |
| Hip function | Harris Hip Score (HHS) |
| Somatic, verbal, social, and cognitive functions | Functional Independence Measure (FIM) |
| Activity of daily living | Modified Barthel index (MBI) |
| Walking ability | KOVAL |
|  | Walking 300 m or more in 6 minutes |
|  | Timed up and Go test |
|  | Walk independently without any aids |
|  | Gait speed (50-ft and 4-m fast walk) |
|  | Short Physical Performance Battery (SPPB) |
| Cognitive functions | Mini mental status examination (MMSE) |
| **Quality of life and wellbeing** |  |
| Quality of life | EuroQoL (EQ-5D-3L) |
| Anxiety | Self-Rating Anxiety Scale scores |
| Depression | Self-Rating Depression Scale scores |
| **Other** |  |
|  | Mortality |
|  | Pain evaluation |
|  | Length of hospital stay |
|  | Rate of readmission and re-operations |
|  | Off-bed time |
|  | Postoperative complications |
|  | Anti-osteoporotic therapy |
|  | Housing |
|  | Satisfaction during hospitalisation |

### Randomised Controlled Studies

The RCT studies were from South Korea (Aftab et al., 2020); USA (Magaziner et al., 2019) and China (Zhang et al., 2022), and all were of high quality as assessed by the quality appraisal RCT checklist (The Joanna Briggs Institute, 2020), but differed with regards to follow-up time and beneficial results.

The RCT study from Korea conducted with hip fracture patients found significant differences between the Fragility Fracture Integrated Rehabilitation Management (FIRM) rehabilitation group and the comparison group on the 15^th^ day post-operation. The patients received 10 FIRM rehabilitation sessions lasting 40 minutes each, with a range of healthcare professionals including physiotherapists, occupational therapists, dieticians, clinical nurses, and social workers. The FIRM group showed significant improvement in stair climbing {0(5) ver. 2(7.5), p=0.049} and ambulation or walker use {8(5) ver. 2(4), p=0.037}, as compared to comparison group (Aftab et al., 2020).

In the USA, a RCT conducted with hip fracture patients found no significant differences between the training group and active control group regarding their ability to walk following hip fracture treatment. At the 16 week follow up and the 40-week follow-up, there was no significant difference in community ambulation between the training group and the active control group who were adults aged 60 years and older with hip fracture (p =.19 at 16 weeks and p=.98 at 40 weeks) (Magaziner et al., 2019).

In China, the RCT conducted with geriatric patients found the timed up and go test (TUG) and short physical performance battery (SPPB) scores in the telerehabilitation group were significantly better than those in the telephone group at 3 months after the intervention, (p < 0.05). The Internet-based rehabilitation management system applied to postoperative home rehabilitation of geriatric hip fractures was found to improve the functional recovery of the hip joint and enhance the ability to perform activities of daily living and somatic integration to a certain extent. In addition, the system is simple to operate and easy to use. For medical workers, the system facilitates the management of basic information and rehabilitation data, and can effectively track the rehabilitation process of patients to achieve personalised rehabilitation treatment (Zhang et al., 2022).

### Cohort studies

The three cohort studies were from Germany, Austria and Switzerland (Schoeneberg et al., 2021); China and Hong Kong (Xiang et al., 2021), and China (Zhong et al., 2021). These studies were of moderate to high quality as appraised by the cohort appraisal checklist (Moola et al., 2017).

A retrospective cohort study from Austria, Germany and Switzerland showed that early geriatric rehabilitation (EGR) led to a significant reduction in mortality (p=0.011) after 120 days in a sample of orthogeriatric patients with hip fracture 4 months after surgery. However, there was also a significant reduction in quality of life (p=0.026). Mortality was reduced as compliance to the anti-osteoporotic medication increased (Schoeneberg et al., 2021).

Another retrospective cohort study from China and Hong Kong, showed that early mobilisation resulted in a significantly better Modified Barthel Index than late mobilisation (mean [SD]: 83.7 [12.0] vs. 67.0 [17.5], p < .001). Adjusted mixed effects model showed significantly higher Modified Barthel Index for early mobilisation at postoperative visit, six weeks, and 12 weeks (all p < .001). Patients in the early mobilization group had slightly better EQ-5D Index at 12 weeks than patients in the late mobilisation group (mean: 0.91 vs 0.87, p = .002), but the difference was small and may not have been clinically relevant (Xiang et al., 2021).

A prospective cohort study conducted in China with 348 patients who had undergone total hip arthroplasty in one hospital in China between 2015 and 2018, found that the application of rapid rehabilitation surgery in total hip arthroplasty can accelerate the postoperative recovery of patients, relieve anxiety and depression, and increase patient satisfaction with the treatment. Compared with the patients in the conventional rehabilitation group, those in the rapid rehabilitation group had shorter hospital stays (11.5±1.2day vs 15.5±2.3day, p = .021), resumed off-bed activities sooner (20.5±3.4hours vs 61.8±4.7 hours, p = 001, had less postoperative pain (4.0±1.2 vs 6.5±1.1, p <.001), and lower anxiety and depression scores (anxiety score: 24.4±2.1 vs 47.9±2.9; depression score: 25.8±1.8 vs 43.7±1.7, p <.001). (Zhong et al., 2021).

## Discussion

The objective of this rapid review was to find recent evidence of clinical effectiveness and economic effectiveness of hip fracture rehabilitation in older people. Hip fracture is a common problem among older adults and with an increase in patients with hip fracture comes and increase in the number of patients who require rehabilitation (Kang et al., 2023).

Six papers were included in this rapid review which focussed on hip fracture rehabilitation programmes published since 2017. The studies were conducted in North America (USA), Europe (Germany, Austria and Switzerland) and Asia (Hong Kong and mainland China) (Aftab et al., 2020; Magaziner et al., 2019; Schoeneberg et al., 2021; Xiang et al., 2021; Zhang et al., 2022; Zhong et al., 2021). Some RCTs and some cohort studies found beneficial effects of rehabilitation for older people who had undergone hip replacement treatments. The length of follow-up period ranged from 15 days to one year after surgery. Two studies reported on quality of life using EuroQoL (EQ-5D) (Schoeneberg et al., 2021; Xiang et al., 2021). Even though it is known that psychological distress, including loss of confidence and frustration, is the most important predictor of depression and anxiety in patients with hip fractures (Ko et al., 2021), only one study assessed the anxiety and depression status of patients after surgery (Zhong et al., 2021). It was not possible to meta analyse the studies as all had different follow-up periods and used varying outcome measures.

Despite the variation in outcome measures, the results indicated that there are beneficial outcomes and a reduction in mortality in cohorts of the elderly population who undergo early rehabilitation following hip fracture treatment (Schoeneberg et al., 2021). The quality of life of people in the early rehabilitation group decreased at 4 months after intervention (Schoeneberg et al., 2021),while another study showed the improvement of EQ-5D index at 12 weeks after intervention (Xiang et al., 2021).

The research by Magaziner et al. (2019) and Aftab et al. (2020) were mentioned in the recent systematic reviews (Fairhall et al., 2022; Handoll et al., 2021). Only one additional RCT was found in our rapid review (Zhang et al., 2022), however we also found three additional cohort studies.

No new economic outcomes for hip fracture have been reported since 2015 in the UK, and only one other from Norway was identified (Taraldsen et al., 2019). The study from Norway investigated the cost-effectiveness of home-based exercise programme after hip surgery (Taraldsen et al., 2019). The original FEMuR study only reported cost outcomes of rehabilitation after hip surgery (N. H. Williams et al., 2016).

Interestingly, the internet-based rehabilitation management system used in China can improve the functional recovery of the hip joint after hip fracture and enhance the ability to perform activities of daily living (Zhang et al., 2022). Other small sample studies showed improvements in walking ability using walkers (Aftab et al., 2020) whilst others showed no improvements in walking ability outcomes despite early rehabilitation (Magaziner et al., 2019).

This rapid review shows that the goal of achieving personalised rehabilitation treatment is possible, and this concept of goal directed personalised rehabilitation should be developed and economically evaluated further.

Internet-based rehabilitation should be compared with face-to-face rehabilitation programmes for clinical effectiveness and cost-effectiveness. It is plausible that internet-based rehabilitation interventions would be the most sustainable and robust in future pandemics where restrictions and lockdown measures are put into place (Genie et al., 2020).

However, none of the included papers contained an economic evaluation aspect and this is a gap in the research evidence base which will be addressed in the on-going FEMuRIII RCT (N. Williams et al., 2020).

### Strengths

A strength of this review is that robust, recent, and relevant papers regarding rehabilitation following hip replacement were found and described.

### Limitations

The main limitations were that none of the included studies conducted a cost-analysis regarding the intervention they described, and no UK based study was included. However, FEMuRIII is an on-going RCT based in the UK exploring the effectiveness and cost-effectiveness of the enhanced rehabilitation programme following surgical repair of proximal femoral fracture in older people compared with usual care (N. Williams et al., 2020).

## Conclusion

The present rapid review found one additional RCT and three additional cohort studies in recent worldwide literature. The results showed that hip rehabilitation interventions can be effective in helping older adults to maintain activities of daily living and mobility following hip fracture treatment. The review not only highlighted the lack of clinical and cost-effectiveness evidence in this area of healthcare, but also highlighted that internet-based programmes may be cost-effective and a robust way forward for rehabilitation treatment across the world having shown to have worked for older people on the Asian continent during the COVID-19 pandemic.

### Recommendations

It is recommended that:

- Longer follow-up periods are needed from the hip rehabilitation treatment to estimate the long-term effectiveness of community or home-based rehabilitation interventions.
- Quality of life and wellbeing should be focused on people with hip fracture after surgery
- Future hip fracture rehabilitation studies should include an economic evaluation aspect.
- Internet-based rehabilitation programmes should be compared against face-to-face rehabilitation programmes in terms of feasibility, acceptability, clinical effectiveness, and cost-effectiveness.

## Data Availability

All data produced in the present work are contained in the manuscript

