## Supplementary material for "Hip fracture rehabilitation in hospital and community settings: A rapid review of the evidence": Supplemental file_15-01-2025.docx

List of Supplementary files

Supplementary file 1 – PICO framework keywords

Supplementary file 2 – Search strategy for FEMuRIII Rapid Review

Supplementary file 3 – Data extraction tables and quality appraisal tables for FEMuRIII Rapid Review

Supplementary File 1 PICO framework Keywords.

The following table shows the Population, Intervention, Comparison and Outcome (PICO) keywords for the FEMuRIII rapid review.

| **Population** | **Intervention** | **Comparison** | **Outcome** | **Economic terms** |
| --- | --- | --- | --- | --- |
| aged | bone* | control | activities of daily living | cost |
| aging | femur | usual practice | bed occupancy | QALY |
| elderly | fracture |  | functional* | utility |
| old* | hip |  | mobility |  |
|  | orthopaedic |  | mortality |  |
|  | rehabilitation programme |  | pain |  |
|  | thigh |  | quality of life |  |

Included studies: Randomised Controlled Trials (RCTs), primary studies including cross-sectional, cohort, longitudinal, and case studies/case reports.

Excluded publications: Editorials, letters to editors, conference abstracts, commentaries, viewpoint papers.

Supplementary File 2 Search strategy for FEMuRIII Rapid Review

The following is the FEMuRIII search strategy via Medline.

**Femur III search strategy via Medline**

| 1. | (Elderly.mp. or Aged/) adj2 femur.mp. |
| --- | --- |
| 2. | (Aging/ or aging.mp.) adj2 femur.mp. |
| 3. | (old.mp. or Aged/) adj2 femur.mp. |
| 4. | 1 or 2 or 3 |
| 5. | Femur/ or femur.mp. |
| 6. | "Bone and Bones"/in [Injuries femur] |
| 7. | Hip Fractures/ or Aged/ or Fractures, Bone/ or Femoral Fractures/ [adj2 femur] |
| 8. | Hip Injuries/ or Osteoarthritis, Hip/ or Arthroplasty, Replacement, Hip/ or Hip Fractures/ or Hip Joint/ or Hip/ |
| 9. | Hospitals, Rehabilitation/ or "Physical and Rehabilitation Medicine"/ or rehabilitation.mp. [adj2 femur] |
| 10. | Thigh/in [Injuries] |
| 11. | 5 or 6 or 7 or 8 or 9 or 10 |
| 12. | "Activities of Daily Living"/ adj2 Femur.mp. |
| 13. | Bed Occupancy/ [adj2 femur] |
| 14. | mobility.mp. or Aged/ [adj2 femur] |
| 15. | Mortality/ [adj2 femur] |
| 16. | Pain/ [adj2 femur] |
| 17. | 12 or 13 or 14 or 15 or 16 |
| 18. | Quality-Adjusted Life Years/ |
| 19. | Aged/ or Cost-Benefit Analysis/ or QALY.mp. or Quality-Adjusted Life Years/ or "Quality of Life"/ |
| 20. | cost.mp. or "Costs and Cost Analysis"/ |
| 21. | economic evaluation.mp. or Cost-Benefit Analysis/ |
| 22. | "Quality of Life"/ or utility.mp. |
| 23. | cost-effectiveness.mp. or Cost-Effectiveness Analysis/ |
| 24. | 18 or 19 or 20 or 21 or 22 or 23 |
| 25. | 4 and 11 and 17 and 24 |
| 26. | 25 and 2017:2023.(sa_year). |

Supplementary File 3 Data extraction tables and quality appraisal tables

The data extraction tables and quality appraisal tables are show below for the randomised controlled (RCT) studies (The Joanna Briggs Institute, 2020), and the cohort studies (Moola et al., 2017). See Tables 1 to 4.

**Table 1: Summary of included Randomised Controlled Study studies for the FEMURIII Rapid Review**

Example of table for primary studies

| **Citation (Country)** | **Study Details** | **Participants & setting** | **Key findings** | **Observations/notes** |
| --- | --- | --- | --- | --- |
| Aftab et al 2020  (Aftab et al., 2020)  **South Korea** | **Study Design:** Randomised Controlled Trial (RCT)  **Related publication: No**  **Type of intervention:**  Fragility fracture integrated rehabilitation management (FIRM) on mobility, activity of daily living and cognitive functioning in elderly with hip fracture.  The FIRM rehabilitation programme.  Each participant was admitted for 15 days after surgery and received 10 physical therapy (PT) sessions/ (FIRM #1-10) and 4 occupational therapy (OT) sessions (FIRM #4, 6, 8, 10). PT sessions included weight-bearing, strengthening, gait training, aerobics, and functional exercises. The duration of each session was at least 40 Minutes. Occupational therapy included training of activities of daily living (ADLs) transfer, sit to stand, bed mobility, dressing, self-care retraining, and use of adaptive equipment. Multidisciplinary rehabilitation members also provided comprehensive patients education. Conventional postoperative rehabilitation involved PT for 40 min/day, ward education, fall prevention, discharge planning, including in-hospital, post- operative usual orthopaedic care. Ward education included techniques about carrying out clothing, carrying out transfer, education about standing exercises, bed exercises and strengthening exercises with elastic band and toileting.  **Data collection methods:** The data was collected through KOVAL for walking ability, Modified Barthel index (MBI) for behaviours related to activities of daily living (ADLS) and mini mental status examination (MMSE) for cognitive functions.  **Follow-up period:** on 2^nd^ postoperative day and after 10th FIRM session on 15^th^ postoperative day.  **Quality rating: High** | **Sample size:** 39 in total. n=20 in the intervention group  n=19 in the conventional physical therapy group  **Participants:** Adults aged 65-95 years old with hip fracture.  **Setting:** Hospital setting in South Korea  **Dates of data collection:** August 2017 – January 2018. | **Primary Findings:**  The post intervention comparison did not show any significant difference (p>0.05) in walking ability, overall activities of daily diving and cognitive functioning. But FIRM group showed significant improvement in stair climbing {0(5) ver. 2(7.5), p=0.049} and ambulation or walker use {8(5) ver. 2(4), p=0.037}, as compared to comparison group.  **Additional Findings:** MMSE improved in the intervention group but not in the comparison group (p < .05). | A randomised controlled trial conducted with hip fracture patients in South Korea found significant differences between the FIRM rehabilitation group and the comparison group on the 15^th^ day post-operation. The patients received 10 FIRM rehabilitation sessions lasting 40 minutes each, with a range of healthcare professionals including physiotherapists, occupational therapists, dieticians, clinical nurses, and social workers. The FIRM group showed significant improvement in stair climbing {0(5) ver. 2(7.5), p=0.049} and ambulation or walker use {8(5) ver. 2(4), p=0.037}, as compared to comparison group |
| **Magaziner et al 2019**  (Magaziner et al., 2019)  **USA** | **Study Design:** Randomised Controlled Trial (RCT)  **Related publication: No**  **Type of intervention:**  Multi-component home-based physical therapy intervention.  The training intervention (active treatment) included aerobic, strength, balance, and functional training. The active control group received transcutaneous electrical nerve stimulation and active range-of-motion exercises. Both groups received 2 to 3 home visits from a physical therapist weekly for 16 weeks; nutritional counselling; and daily vitamin D (2000 IU), calcium (600 mg), and multivitamins.  **Data collection methods:** 60-minute home intervention visits on non-consecutive days for 16 weeks. The primary outcome (community ambulation) was defined as walking 300 m or more in 6 minutes.  **Follow-up period:** 16 and 40 weeks after randomization.  **Quality rating: High** | **Sample size:** 210 (n=105 in the Training Group arm, and n=105 in the Active Control Group).  **Participants:** Adults aged 60 years and older with hip fracture.  **Setting:** Home settings in the USA  **Dates of data collection:**  September 2016-October 2017. | **Primary Findings:** At the 16 week follow up and the 40-week follow-up, there was no significant difference in community ambulation between the training group and the active control group (p =.19 at 16 weeks and p=.98 at 40 weeks).  **Additional Findings:** There were improvements in ambulation between baseline and follow-up for both groups, however no differences between groups. | A randomised controlled trial conducted with patients in the USA found no significant differences between the training group and active control group regarding their ability to walk following hip fracture treatment. At the 16 week follow up and the 40-week follow-up, there was no significant difference in community ambulation between the training group and the active control group (p =.19 at 16 weeks and p=.98 at 40 weeks). |
| Zhang et al 2022  (Zhang et al., 2022)  **China** | **Study Design:** Randomised Controlled Trial (RCT)  **Related publication: No**  **Type of intervention:**  Telephone intervention versus telerehabilitation intervention. Patients in both groups received routine discharge instructions 1–2 days after surgery: bedside rehabilitation instructions and health education, including ankle pump exercises, heel gliding exercises, quadriceps contraction training, breathing exercises, and prevention of postoperative complications, dietary instructions, etc. The Telephone Group The group received telephone follow-up after discharge: the patients received telephone follow-ups at 2 weeks, 1 month, 2 months, and 3 months after discharge from the hospital. The patients in the telerehabilitation group were discharged from the hospital, and a “home-oriented post-operative rehabilitation management system for geriatric hip fractures” developed by our department, was used for home-based telerehabilitation. The system consisted of a physician side— a website platform, and a patient side—an application installed on a smartphone.  **Data collection methods:** Patients in both groups received routine discharge instructions 1–2 days after surgery. The Harris hip score (HHS), functional independence measure (FIM), timed up-and-go test (TUG), and short physical performance battery (SPPB) were used to evaluate the patients’ hip function, activities of daily living, and overall somatic ability.  **Follow-up period**: 1 month and 3 months after surgery  **Quality rating: High** | **Sample size:**  n=58 elderly postoperative hip fracture patients (in total).  n=29 in the telephone group.  n=29 in the telerehabilitation group.  **Participants:** Adults aged 60 years and older with hip fracture  **Setting:** Home settings in China.  **Dates of data collection:**  Data was collected between June 2020 and November 2020. | **Primary Findings:** There was no significant difference between the baseline data of the two groups before the intervention (p > 0.05); no matter after hip replacement or internal fixation, the HHS score and FIM score of both groups increased gradually with the postoperative time, and the scores in the telerehabilitation group were higher than those in the telephone group at 1 and 3 months after the intervention, and the difference was significant (p < 0.05); for patients after hip replacement, the TUG and SPPB scores in the telerehabilitation group were better than those in the telephone group at 3 months after the intervention, and the difference was significant (p < 0.05).  **Additional Findings:** The Internet-based rehabilitation management system applied to postoperative home rehabilitation of geriatric hip fractures can improve the functional recovery of the hip joint and enhance the ability to perform activities of daily living and somatic integration to a certain extent. In addition, the system is simple to operate and easy to use. For medical workers, the system facilitates the management of basic information and rehabilitation data and can effectively track the rehabilitation process of patients to achieve personalised rehabilitation treatment. The system can also automatically deliver rehabilitation videos according to the patient’s condition, which can reduce the clinical workload to a certain extent. | A randomised controlled trial conducted with patients in China found TUG and SPPB scores in the telerehabilitation group were significantly better than those in the telephone group at 3 months after the intervention, (p < 0.05). |

**Abbreviations:** Randomised Controlled Trial (RCT).

See Table 2 for the JBI RCT quality appraisal table.

**Table 2 JBI critical appraisal checklist for randomised controlled trials** (The Joanna Briggs Institute, 2020)**.**

| **Study** | **JBI Appraisal items** | | | | | | | | | | | | | **Score** |
| --- | --- | --- | --- | --- | --- | --- | --- | --- | --- | --- | --- | --- | --- | --- |
|  | **Q1** | **Q2** | **Q3** | **Q4** | **Q5** | **Q6** | **Q7** | **Q8** | **Q9** | **Q10** | **Q11** | **Q12** | **Q13** |  |
| Aftab et al 2020 | Y | N | Y | N | N | U | Y | Y | Y | Y | Y | Y | Y | High |
| Magaziner et al 2019 | Y | Y | Y | N | N | U | U | Y | Y | Y | Y | Y | Y | High |
| Zhang et al 2022 | Y | N | Y | N | N | U | U | Y | Y | Y | Y | Y | Y | High |

Key: Y – Yes; N – No; U – Unclear; n/a – not applicable

1. Was true randomization used for assignment of participants to treatment groups?
2. Was allocation to treatment groups concealed?
3. Were treatment groups similar at the baseline?
4. Were participants blind to treatment assignment?
5. Were those delivering treatment blind to treatment assignment?
6. Were outcomes assessors blind to treatment assignment?
7. Were treatment groups treated identically other than the intervention of interest?
8. Was follow up complete and if not, were differences between groups in terms of their follow up adequately described and analysed?
9. Were participants analysed in the groups to which they were randomized?
10. Were outcomes measured in the same way for treatment groups?
11. Were outcomes measured in a reliable way
12. Was appropriate statistical analysis used?
13. Was the trial design appropriate, and any deviations from the standard RCT design (individual randomization, parallel groups) accounted for in the conduct and analysis of the trial?

**Table 3: Summary of included Cohort studies for the FEMURIII Rapid Review**

| **Citation (Country)** | **Study Details** | **Participants & setting** | **Key findings** | **Observations/notes** |
| --- | --- | --- | --- | --- |
| Schoenberg et al 2021  (Schoeneberg et al., 2021)  **Germany**  **Switzerland and Austria** | **Study Design:** A retrospective cohort study  **Related publication: No**  **Type of intervention:** The German Trauma Society (DGU) began developing the structures of orthogeriatric co-management in cooperation with the geriatric professional association which early geriatric rehabilitation (EGR) is a sub-model of DGU.  **Data collection methods:** Retrospective data four months after surgery. The primary outcomes are the rate of readmission, rate of re-surgery, anti-osteoporotic therapy, housing, mortality, walking ability, and quality of life (EQ-5D-3L).  **Follow-up period:** 120 days post-surgery  **Quality rating: Moderate** | **Sample size:** Total n= 9,780  Early geriatric rehabilitation (EGR) group n = 7,066.  **Participants:** patients with proximal femur fracture who aged 70 years and older. Orthogeriatric patients with hip fracture 4 months after surgery.  **Setting:** Data was obtained from the Registry for Geriatric Trauma DGU (ATR-DGU) from 2016 through 2019, about 25,000 cases from 100 centres in Germany, Switzerland, and Austria.  **Dates of data collection:** 2016-2019 (only centres in Germany were included in the analysis of EGR) The total sample also included centres from Switzerland and Austria). | **Primary Findings:** The Early geriatric rehabilitation (EGR) showed a positive influence of anti-osteoporotic treatment (p<0.001) and mortality (p=0.011) but led to a slight reduction in QoL (p=0.026).  **Additional Findings:** The EGR has a significant impact on compliance in the prescription of anti-osteoporotic medication and leads to a significant reduction in mortality after 120 days. | A retrospective cohort study from Austria, Germany and Switzerland showed that early geriatric rehabilitation (EGR) led to a significant reduction in mortality (p=0.011) after 120 days, but also a significant reduction in quality of life (p=0.026). Mortality was reduced as compliance to the anti-osteoporotic medication increased. |
| Xiang et al 2021  (Xiang et al., 2021)  **China and Hong Kong, China** | **Study Design:** multicentre prospective cohort study.  **Related publication: No**  **Type of intervention:**  Early mobilization versus late mobilization after intertrochanteric fracture treatment. Early mobilization scheme was defined as transferring from bed to a sitting chair within 2 days after surgery, standing up with both feet on the ground within 4 (±2) day after surgery, and walking starting within 5 (±2) day after surgery. Late mobilization scheme was defined as weight-bearing walking (with or without walking aids) starting more than 7 days after surgery as per local standard. Patients in both groups performed immediate in-bed mobilization after surgery. After discharge, all patients followed a standardized daily exercise program at home during the first 12 weeks.  **Data collection methods:** All patients performed immediate in-bed mobilization after surgery and followed a standardized  daily exercise program at home during the first 12 weeks. Functional status was measured by the Modified Barthel Index at  postoperative visit, 6 weeks, and 12 weeks. QoL was measured by the EuroQol-5D (EQ-5D) at 12 weeks. The total follow-up time was 1 year from the day of surgery. The postoperative visit was performed 4 (±2) days after the surgery.  **Follow-up period:** Patients attended clinic follow-up visits at 6 and 12 weeks. The 1-year visit was conducted via a telephone interview.  **Quality rating: High** | **Sample** **size:**  n = 148 were enrolled to early mobilization, and  n = 136 to late mobilization.  **Participants:** Patients (≥65 years old) with unstable intertrochanteric fractures treated with intramedullary nails were recruited from nine centres in China.  **Setting:** Hospital settings in China.  **Dates of data collection:** Data was collected between April 2015 and April 2017. | **Primary Findings:** At 6 weeks, early mobilization resulted in a significantly better Modified Barthel Index than late mobilization (mean [SD]: 83.7 [12.0] vs. 67.0 [17.5], p < .001). Adjusted mixed effects model showed significantly higher Modified Barthel Index for early mobilization at postoperative visit, 6 weeks, and 12 weeks (all p < .001). Patients in the early mobilization group had slightly better EQ-5D Index at 12 weeks than patients in the late mobilization group (mean: 0.91 vs 0.87, p = .002).  **Additional Findings:** Early postoperative mobilization resulted in better functional outcomes up to 12 weeks. QoL was rated statistically significantly better in the early mobilization group, but the difference was small and may not be clinically relevant. | A retrospective cohort study from China and Hong Kong, China showed that early mobilisation resulted in a significantly better Modified Barthel Index than late mobilization (mean [SD]: 83.7 [12.0] vs. 67.0 [17.5], p < .001). Adjusted mixed effects model showed significantly higher Modified Barthel Index for early mobilization at postoperative visit, 6 weeks, and 12 weeks (all p < .001). Patients in the early mobilization group had slightly better EQ-5D Index at 12 weeks than patients in the late mobilization group (mean: 0.91 vs 0.87, p = .002), but the difference was small and may not be clinically relevant. |
| Zhong et al 2021  (Zhong et al., 2021)  **China** | **Study Design:** A prospective cohort study.  **Related publication: No**  **Type of intervention:**  Early postoperative exercises: The patients started simple exercises such as quadriceps static contraction and ankle dorsiflexion 6 hours after the operation. Urinary tubes and incision drainage tubes were withdrawn within 2 days after the operation. Patients could stretch the operated leg and do lifting exercises. Within 1 week after surgery, patients moved from lying to standing gradually and to walk without crutches. The rehabilitation training required a rehabilitation doctor’s professional guidance to help patients stop using crutches and walk on their own as soon as possible.  Conventional rehabilitation methods: before surgery, the patients received only education regularly provided on admission. The patients gradually began rehabilitation exercises and walked without weight-bearing at approximately 14 days after surgery.  **Data collection methods:** The following outcomes were measured:   - Length of hospital stay - Time to off-bed activity - Pain score - Self-Rating Anxiety Scale scores - Self-Rating Depression Scale scores - Complication rate - Rate of satisfaction during hospitalization.   **Follow-up period**: within one year after surgery  **Quality rating: High** | **Sample size:** N=348 in total.  N=180 received rapid rehabilitation nursing  N=168 patients received conventional nursing.  **Participants:** 348 people who had undergone total hip arthroplasty at one hospital in China. 58< patients<90 years of age.  **Setting:** Hospital setting in China.  **Dates of data collection:**  January 2015 to December 2018. | **Primary Findings:** Compared with the patients in the conventional rehabilitation group, those in the rapid rehabilitation group had shorter hospital stays (11.5±1.2day vs 15.5±2.3day, p=.021), resumed off-bed activities sooner (20.5±3.4hours vs 61.8±4.7 hours, p=.001, had less postoperative pain (4.0±1.2 vs 6.5±1.1, p<.001), and lower anxiety and depression scores (anxiety score: 24.4±2.1 vs 47.9±2.9; depression score: 25.8±1.8 vs 43.7±1.7, p<.001).  **Additional Findings:** Rapid rehabilitation surgery can reduce depression and anxiety in patients with total hip arthroplasty. | A prospective cohort study was conducted in China with 348 patients who had undergone total hip arthroplasty in one hospital in China between 2015 and 2018. It was found that the application of rapid rehabilitation surgery in total hip arthroplasty can accelerate patients’ postoperative recovery, relieve anxiety and depression, and increase the patient’s satisfaction with the treatment. |

See Table 4 for the JBI cohort study quality appraisal.

**Table 4 JBI critical appraisal checklist for cohort studies (prospective)** (Moola et al., 2017)

| **Study** | **JBI Appraisal items** | | | | | | | | | | | **Score** |
| --- | --- | --- | --- | --- | --- | --- | --- | --- | --- | --- | --- | --- |
|  | **1** | **2** | **3** | **4** | **5** | **6** | **7** | **8** | **9** | **10** | **11** |  |
| Schoenberg et al 2021 | N | N | Y | Y | Y | Y | Y | N | Y | Y | Y | Moderate |
| Xiang et al 2021 | Y | Y | Y | Y | Y | Y | Y | Y | Y | Y | Y | High |
| Zhong et al 2023 | Y | Y | Y | Y | Y | Y | Y | Y | Y | Y | Y | High |

Key: Y: Yes; N: No; U: Unclear; n/a: not applicable

1. Were the two groups similar and recruited from the same population?
2. Were the exposures measured similarly to assign people to both exposed and unexposed groups?
3. Was the exposure measured in a valid and reliable way?
4. Were confounding factors identified?
5. Were strategies to deal with confounding factors stated?
6. Were the groups/participants free of the outcome at the start of the study (or at the moment of exposure)?
7. Were the outcomes measured in a valid and reliable way?
8. Was the follow up time reported and sufficient to be long enough for outcomes to occur?
9. Was follow up complete, and if not, were the reasons to loss to follow up described and explored?
10. Were strategies to address incomplete follow up utilized?
11. Was appropriate statistical analysis used?
